# Culture and understanding the role of feedback for health professions students: Realist synthesis protocol

**DOI:** 10.1101/2021.01.24.21250413

**Authors:** Paul Fullerton, Mahbub Sarkar, Shamsul Haque, Wendy McKenzie

**Affiliations:** Jeffrey Cheah School Medicine & Health Sciences, Monash University Malaysia; Monash Centre for Scholarship in Health Education, Monash University, Clayton, VIC 3800, Australia; Jeffrey Cheah School Medicine & Health Sciences, Monash University Malaysia, Bandar Sunway, Selangor Malaysia; Education, Faculty of Medicine, Nursing & Health Sciences, Monash University, Clayton, VIC 3800, Australia

**Keywords:** Realist review, Realist Synthesis, Health professional students, Medical students, Feedback, Asia, Protocol

## Abstract

**Introduction:** Clinical education has moved to a “competency-based” model with an emphasis on workplace-based learning and assessment which, in turn, depends on feedback to be effective. Further, the understanding of feedback has changed from information about a performance directed to the learner performing the task, to a dialogue, which enables the learner to act and develop.

In health professional education, feedback is a complex interaction between trainee, supervisor, and the healthcare system. Most published research on feedback in health professional education originates in Europe and North America. Our interest is on the impact of **C**ulture on this process, particularly in the context of Asian cultures.

The (scientific) realist approach of Pawson and Tilley provides a means to examine complex interventions in social situations, and thus is an appropriate lens to use for this study. This is a protocol for a realist synthesis which asks how, why and in what circumstances do Asian Cultures influence health professional trainees to seek, respond to and use feedback given in the clinical environment, if at all.

**Methods and analysis:** An initial search was performed to help define the scope of the review question and develop our initial program theory. The formal electronic search was carried out in February 2020 and included: CINAHL, ERIC, MEDLINE, and PsycInfo, and repeated in October 2020.

Retrieved articles were imported into Covidence for screening and data extraction, after which components of the Context – Mechanisms – Outcomes configurations will be sought to refine the initial program theory.

**Ethics and Dissemination:** As this study is a literature review, ethics approval is not required.

The findings will be documented in line with the RAMESES publications standards for Realist syntheses,[41] and we plan to disseminate the findings by means of a peer-reviewed journal article and conference presentation(s).

**Strengths and limitations of this study:**

- The synthesis aims to identify the how and why Asian **C**ultures may influence feedback seeking and provision to health professional trainees, if at all.
- To our knowledge, there are few studies of feedback seeking and provision to health professional trainees in Asia.
- A Realist approach has the potential to help explain the complex nature of **C**ulture’s impact on feedback.
- Only studies published in the English language will be included, so transferability of our findings to non-English speaking environments may be lacking.
- In addition to formal literature database searches, we will need to conduct citation mining to locate other relevant resources.

## INTRODUCTION

Clinical education has moved to a “competency-based” model with an emphasis on workplace-based learning and assessment which, in turn, depends on feedback to be effective.[1,2] Indeed Ramani *et al*[2](p744) describe feedback as “*a vital cog in the wheel of competency-based medical education*.” Given that feedback is established as an important link in competency-based medical education, this led to our interest in the impact of **C**ulture on the feedback process, understanding of the tools used, and its acceptance by both supervisors and trainees.

### Complexity of Feedback

Early definitions of feedback emphasised information giving to change behaviour.[3–5] In a widely quoted paper, Ende[3] described feedback as ***information*** given to trainees about a particular activity which was meant to guide performance of that or a similar activity in the future. The emphasis was that feedback was something supervisors directed at trainees, preferably after observation of the activity in question. University students commonly complain that they do not receive enough feedback, or that it is done poorly, such that academic staff are advised to “signpost” when feedback was being given.[6] Ajjawi and Regehr[7] suggest that perhaps learners and teachers define feedback quite differently.

Over time, feedback has been understood as more than simply providing information – information is only feedback when it is ***used*** to improve work or learning and is part of a sociocultural interaction. Furthermore, feedback value is influenced by the credibility of the feedback source.[8–10] As it is essential to close the feedback loop, we can think of feedback as sense-making in the context of information provided from many sources to improve work and learning.[11] The importance of relationships and trust between the supervisor and trainee, especially when the feedback relates to assessment, has been emphasised.[12] Many factors influence the effect of feedback including context (e.g. the workplace – hospital or ambulatory settings, teaching a skill, formative assessment, summative assessment)[13,14] regulatory focus,[15] and self-efficacy.[16,17] A person’s “theory of intelligence” (their understanding of whether intelligence is “fixed” or “improvable”) will also impact – if a person’s belief is that intelligence is fixed, effort may not seem to be worthwhile, whereas if they feel there is opportunity for improvement, effort becomes worthwhile.[18,19]

Feedback within the clinical learning environment is particularly complex and influenced by such things as the workload of providing patient care, hierarchies, time constraints and limited opportunities to observe a student’s performance, the student’s expectations and engagement with feedback provided,[8,16,20] as well as the supervisor’s experience of feedback during their training and therefore understanding of feedback. These complexities will be recognised by clinical teachers in Western environments, but we suspect are magnified within the Asian setting.[21]

### Complexity of Culture

When we consider culture in the context of health professional education there are three prominent and interdependent cultures – the “big-**C**ulture”, the Workplace culture and the Education culture.

> **C**ulture (sometimes referred to as “big-Culture”) in this context is defined by Hofstede as *“The collective programming of the mind that distinguishes one group or category of people from another* … *culture is (a) a collective, not individual, attribute; (b) not directly visible but manifested in behaviours; and (c) common to some but not all people*.*”*. [22](p58)

While there are several classifications of characteristics of **C**ulture, Hofstede’s typology is widely used and can help in our understanding of the issues. Initially four dimensions were described,[23,24] with two further dimensions added later:[22,25]

1. Individualism – Collectivism
2. Power Distance
3. Uncertainty Avoidance
4. Masculinity – Femininity.
5. Long-term versus Short-term Orientation
6. Indulgence versus Restraint

Of these dimensions, Individualism-Collectivism and Power Distance appear to be the most significant big-**C**ulture influences in the clinical learning environment in South-East Asia.[21]

All authors have experience teaching in health professions education in Asia and Australia in a University with campuses in both Malaysia and Australia. This experience raised the question whether cultural differences may influence our students’ learning. A preliminary literature review of feedback within health professional education showed a heavy North American and European focus, and few studies from a South East Asian perspective, except notably from Indonesia.[21,26] This heightened our questioning of *whether* cultural factors influence acceptance and engagement with “dialogic feedback” in an Asian context, and how does it compare with the “Western” situation? (While some Western studies have included international students / trainees, we contend that students who reached the clinical phase of training have had time and opportunity to adapt to their host country.) In focussing on the Asian region, we can recognise several *broad* **C**ultural groups – the “Confucian Heritage Culture”, Indonesian-Malaysian / Muslim cultures, and cultures of the Indian subcontinent, overlaid with the cultural impacts of colonialism. Given the limited literature found in our preliminary search focussing on Southeast Asia, we decided to look further afield to include the Middle East (the influence of Muslim learning culture) through to the “Far East” (the “Confucian Heritage Culture”). If the cultural background of students / trainees and their teachers / supervisors influences their engagement with feedback, how does it do so? Is it the “ethnic **C**ulture”, the national cultures influenced by their colonial history (e.g. for Malaysia, Singapore, Indonesia)[27], or is it the education system culture (e.g. school education, university, or even discipline cultures) that have the predominant effect – or is there no dominant effect?

Workplace culture can be viewed as another cultural layer, particularly in clinical teaching and hospital environments[28] and potentially interacts with the “big-**C**ulture”. Medicine around the world tends to be hierarchical and paternalistic. However, within the Asian region teaching by humiliation is common, and in Malaysia the term “scolding” is commonly used to describe teaching in the clinical environment. Another term often heard in the region is *kiasu* – particularly applied to students of Chinese ethnicity. According to the Oxford Dictionary,[29] *kiasu* refers to a person who is *“governed by self-interest, typically manifesting as a selfish, grasping attitude arising from a fear of missing out on something*.*”. Kiasu* (怕输) is a Hokkien word meaning “fear of loss”. There are two aspects to *kiasu*, the negative side of being selfish and grasping, as seen in the Oxford dictionary definition, but there is the positive aspect of being successful through hard work – not evident in that dictionary definition.[30] (*Kiasu* is related to the concept of “face”, which western stereotypes frequently regard as a characteristic of “Asian Culture”).

The education system clearly has its overarching culture which influenced the students’ experiences of school – a system that emphasises regular high-stakes examinations from an early stage of schooling. There has been much written about the influence of the “Confucian Heritage Culture” of learning (CHC) on students from South East Asia as well as from China, Japan and Korea. In his writings Confucius saw learning as a means of social change and to overcome social differences, but also placed much emphasis on personal effort.[31] The Chinese philosophy of education also highlighted a mutually respectful relationship between teacher and learner, with the teacher guiding the learner, rather than pulling the learner along.[32] This parallels the role of *guru* seen in the Indian culture of education – with the *guru* (teacher) nurturing the learner.[33,34]

Malaysia and Indonesia are predominantly Islamic countries; Malaysia is a former British colony, while Indonesia was formerly colonised by the Dutch. Clearly both these aspects of their history have shaped the education system of the respective country and are as important factors as the Confucian heritage. There is a great diversity in Islamic education which impacts the southeast Asian region and interacted with the colonial experiences.[35] The school culture clearly responds to the education system’s overarching culture but adds its own layer. An examination-oriented curriculum was seen as a legacy of the colonial era.[36]

Tertiary education culture varies enormously across the region, from hierarchical approaches to being more collegial (especially in the later stages of the degrees). Student experiences in high school impact their transition to university as they come with an expectation that university would simply be an extension of school – first year medical students in Malaysia clearly started university with the idea that knowledge was fixed and largely unchanging, and that their teachers or lecturers functioned as sources of knowledge who were not to be questioned. Knowledge was facts, and facts were immutable.[37] As school had emphasised rote learning of fixed knowledge, and an important part of those students’ adaptation to university was coming to terms with thinking for themselves. As they move into workplace-based learning, the culture of the medical workplace is likely to have an impact.

## METHODS AND ANALYSIS

### Realist Synthesis methodology

The (scientific) Realist approach was chosen as a methodology that is useful for researching complex interventions in the social environment, such as healthcare and education – interventions that frequently work differently in different environments.[38] The more “traditional” methods used in reviews in Medicine and other Health Sciences (such as Systematic Reviews) were felt to not capture or explain the complexities of feedback in the social and cultural environment. The realist paradigm asks: *“What is it about this intervention that works, for whom, in what circumstances, in what respects and how?”*. [39,40] It seeks to find “mechanisms (M)” that fire in particular “contexts (C)” to produce the “Outcomes (O)” in question – so called CMO Configurations. Realist synthesis or realist review (the terms are used interchangeably) is a theory driven, iterative and explanation-building approach, that usually starts with an initial program theory and uses findings from sources to understand how and why the outcomes have occurred, and therefore refine the initial program theory.[38] Interpretation involves looking for both confirming and negating data and explanations.

This synthesis asks what leads can **C**ulture provide, in the Asian health professional education environment, to answer?

1. How, why and in what circumstances do ***health professional trainees*** (e.g. students and junior doctors) seek, respond to, and use feedback given in the clinical environment?
2. What do ***supervisors*** (e.g. consultants, clinical tutors, preceptors) feel about providing feedback? How do they provide feedback, in what circumstances? Do they see their feedback being used?
3. How do ***trainees and supervisors*** perceive feedback?

The review will follow the five steps of a realist review as enunciated by Pawson and colleagues,[39] namely

1. Clarify the scope and purpose of the review question
2. Search for evidence – commencing with an exploratory search, with subsequent focussing and purposive and “snowball” sampling
3. Appraise studies and extract data
4. Synthesise the evidence to obtain conclusions, and …
5. Disseminate.

In appraising studies, ***Relevance*** is assessed by whether it can contribute to theory building or testing, while ***Rigor*** assessment is based on whether the methods which generated a *particular piece of data* is trustworthy.[41] Pawson argues that the overall methodological quality of a study is not appropriate grounds for excluding a study in realist reviews *– “There are often nuggets of wisdom in methodologically weak studies”*.[42]

A PRISMA-P checklist has been completed and available as an additional file.[43]

### Search Strategy

An preliminary search was performed to define the scope of the review question and develop our candidate initial program theory (IPT). This first search utilised MEDLINE and PsycInfo, searching “Learner” (and variations), Feedback (and debrief) and Culture (including cross-cultural, ethnic differences, anthropology). In terms of a modified PICo model[44] developed for Qualitative studies (**P**opulation, **I**ntervention, **Co**ntext) format: P: Learners, I: Feedback, Co: Culture.

The formal electronic search was carried out in February 2020 and included: CINAHL, ERIC, MEDLINE, and PsycInfo. Search terms were developed in discussion with a librarian and the research team, with the same broad categories as before, although only articles available in English were retained. Articles to be considered include qualitative, quantitative and mixed methods, as well as commentaries and review articles. An example of the search strategy is given in *Table 1* and provided in more detail as a supplementary file. Both MeSH (medical subject headings) and free text were employed to ensure sufficiently wide article coverage. This search was repeated in October 2020, for articles published since the February search. A hand search will also be made of the following journals: Academic Medicine, Medical Education, Medical Teacher, BMC Medical Education, Education for Health, Teaching and Learning in Medicine, Perspectives on Medical Education, Medical Journal of Malaysia, Annals of the Academy of Medicine Singapore, and Singapore Medical Journal. These were chosen as leading health professional education journals and Southeast Asian medical journals published in English. Citation mining (“snowball”) searches of the reference list in included articles and searching for articles that cite these articles will occur. Although dissertations were initially excluded, relevant published articles arising from the dissertations will be sought by hand-searching for author and a related title.

**Table 1:** Example of Search strategy used in Ovid MEDLINE

| Population: **Learner** | Intervention: **Feedback** | Context: **Culture** |
| --- | --- | --- |
| Subject Headings e.g. MeSH | Subject Headings e.g. MeSH | Subject Headings e.g. MeSH |
| Students, Health Occupations<br>Clinical clerkship<br>Education, medical/... nursing/... pharmacy/ ... public health professional<br>Clinical competence<br>Faculty<br>Faculty, dental/... medical/... nursing | FORMATIVE FEEDBACK<br>Debrief | CULTURE<br>Cross-Cultural Comparison<br>Cultural diversity<br>Cultural difference |

**Table 1:** Example of Search strategy used in Ovid MEDLINE
| Keywords and phrases | Keywords and phrases | Keywords and phrases |
| --- | --- | --- |
| Trainee<br>Student<br>Learner<br>Graduate<br>Intern<br>Supervisor<br>Teacher<br>Lecturer<br>Instructor<br>Professor<br>Tutor | Feedback<br>Feeding back<br>Feed-back<br>Feedforward<br>Feed forward<br>Feeding forward<br>Fed back<br>Debrief | Culture<br>Cultural difference<br>Cultural diversity<br>Cultural understanding<br>Cross cultural<br>Ethnic |

### Selection criteria

Following the searches as outlined, the citations were imported into Covidence[45] for Title and Abstract Screening. Duplicates were removed before title and abstract screening began with two team members (PDF and MS) reviewing a sample of the articles retrieved to ensure that criteria are agreed upon. Approximately 10% of retrieved articles were reviewed jointly by the two team members. The rest of this phase was carried out by either of those team members (predominantly PDF) but with the intention to err on retaining studies for closer evaluation at the full text screening stage.

<u>Core inclusion</u> criteria sought studies relating to

- Workplace-based learning and assessment,
- Feedback giving, seeking and acceptance,
- Culture (Ethnic and institutional),
- Post-secondary and vocational education involving health professional training.

<u>Exclusion</u> criteria centre around:

- Not related to health professional education
- Not related to feedback or culture
- Community health education

Following title and abstract screening, the full texts of the retained articles were imported into Covidence for further screening. All full text articles will be screened by two members of the team, and any discrepancies will be discussed to resolve the disagreements. Notes will be made to justify inclusion or exclusion and will assist with both resolving discrepancies and providing transparency. Selecting papers for the review will be guided by the research study questions – Does the study involve students in health professional courses (especially in their clinical training) or their supervisors? Does the study pertain to students in Asian countries?

Studies remaining after the full text screen will be assessed for Quality and Rigour using the Critical Appraisal Skills Programme (CASP-Qualitative) checklist for Qualitative studies[46] and the Medical Education Research Study Quality Instrument (MERSQI) for quantitative studies.[47] Each study will be appraised by at least two team members – usually PDF and one other. Any discrepancies will be resolved by the full team.

### Extracting Data

Data extraction will follow, with data entered into a table within Covidence 2.0. Data extracted will include citation details, country or region where study was performed, population studied, methodology used, and an empirical judgement of Realist Relevance will also be made at this stage. Comments of potential context, mechanism and outcome will be recorded. Data extracted in Covidence will be exported as a .csv file into Excel, to be used in the synthesis phase. Finally, included articles will be entered into NVivo software[48] for further data extraction, coding and identification of CMO configurations.

### Synthesise findings to draw conclusions

NVivo software in conjunction with the Excel spreadsheet exported from Covidence will be used to synthesise findings and modify our initial program theory. The realist approach involves looking for causal mechanisms and how they fire in particular contexts to produce their outcome(s). Data will be extracted from the included papers by the team of reviewers, with a minimum of 10% of papers being double checked. The data will be tabulated and potential Contexts (C), Mechanisms (M) and Outcomes (O) identified. Discussion among the team members, with the use of realist logic, will aim to further refine those Contexts (C), Mechanisms (M) and find Outcomes patterns (O) to refine our initial program theory and infer CMO configurations. This process will look for confirming and contradictory findings and will be iterative.

### Potential limitations of the realist synthesis approach

We accept that there are possible limitations of this proposed realist synthesis. Firstly, we have confined our search to trainees in clinical training for health profession disciplines, thus limiting the generalisability outside this sphere of education. Secondly, our initial interest was in the Southeast Asian region but due to paucity of literature from SE Asia we expanded the geographical scope. However, the is significant overlay of **C**ultures between the Middle East with the impact of Islam, through to the Far East with the influence of the Confucian Heritage Culture, as well as the influence of the various colonising powers. Thirdly, we have decided to assess rigour in our screening of articles, but recognise that there is a debate among realist scholars as to the validity of assessing rigour of a whole study – Pawson emphasises that “nuggets of wisdom” may be found in studies that are methodologically weak.[42]

### Ethics and Dissemination

As this study is a literature review, ethics approval is not required.

## IMPLICATIONS

Feedback has been recognised by others, and recognised by us, as critically important in competency-based health professional education, yet feedback is a complex, socially based “intervention”. Most of the published literature on feedback originates from “Western” cultures. There is reason to expect that components of culture – “big-Culture”, workplace culture and education system cultures will impact the provision, acceptability and use of feedback. Again, complex interactions come into play. The realist approach is a relevant way to examine these processes. This protocol and resulting realist synthesis will inform a planned study which aims to provide further information that may lead to improving the usefulness of feedback within the Malaysian context and hopefully will be relevant in the wider Southeast Asian region.

### Systematic Review Registration

The protocol for this review was judged to be ineligible for registration with the International Prospective Register for Systematic Reviews (PROSPERO), as it did not*” have a direct and clinically-relevant health-related outcome”*.

## Supporting information

Supplementary File-Lit Review Search Strategy

Supplementary File-PRISMA-P Checklist

## Data Availability

Protocol - no data availability

## Author Contributions

PF developed the original idea for this review protocol in collaboration with MS, WMcK and SH. PF wrote the original draft of the protocol, which was subsequently refined by MS, WMcK, SH, and PF. All authors agree to be accountable for all aspects of this protocol. PF is guarantor of the protocol.

## Competing interests

All authors declare that they have no competing interests.

## Funding

This research received no specific grant from any funding agency in the public, commercial or not-for-profit sectors.

## Ethics

Not applicable.

## Patient and Public Involvement

None – Literature review.

## Notes

### Competing Interest Statement

The authors have declared no competing interest.

### Clinical Trial

Not a clinical trial

### Author Declarations

Ethics approval not required - literature review protocol.

### Summary of Updates

This version of the manuscript has been revised in line with reviewer comments - in particular, the original title was shortened. There were comments that aspects were repetitive, and the repeated parts were modified or removed. A reviewer suggested using a PICO structure for the search strategy - thus a modified PICo model developed for Qualitative studies (Population, Intervention, Context) was used.

## References

1 Norcini J. The power of feedback. Med Educ 2010;44:16–7. doi:10.1111/j.1365-2923.2009.03542.x

2 Ramani S, Könings KD, Ginsburg S, et al. Feedback Redefined: Principles and Practice. J Gen Intern Med 2019;34:744–9. doi:10.1007/s11606-019-04874-2

3 Ende J. Feedback in Clinical Medical Education. JAMA J Am Med Assoc 1983;250:777. doi:10.1001/jama.1983.03340060055026

4 Butler DL, Winne PH. Feedback and Self-Regulated Learning: A Theoretical Synthesis. Rev Educ Res 1995;65:245–81. doi:10.3102/00346543065003245

5 Hattie J, Timperley H. The Power of Feedback. Rev Educ Res 2007;77:81–112. doi:10.3102/003465430298487

6 Boud D, Molloy E. What is the problem with feedback? In: Feedback in Higher and Professional Education. 2012. 1–10.

7 Ajjawi R, Regehr G. When I say … feedback. Med Educ 2018;:3–5. doi:10.1111/medu.13746

8 Watling CJ, Driessen E, Van der Vleuten CPM, et al. Beyond individualism: professional culture and its influence on feedback. Med Educ 2013;47:585–94. doi:10.1111/medu.12150

9 Watling CJ. Unfulfilled promise, untapped potential: Feedback at the crossroads. Med Teach 2014;36:692–7. doi:10.3109/0142159X.2014.889812

10 Wilbur K, BenSmail N, Ahkter S. Student feedback experiences in a cross-border medical education curriculum. Int J Med Educ 2019;10:98–105. doi:10.5116/ijme.5ce1.149f

11 Carless D, Boud D. The development of student feedback literacy: enabling uptake of feedback. Assess Eval High Educ 2018;43:1315–25. doi:10.1080/02602938.2018.1463354

12 Carless D. Trust, distrust and their impact on assessment reform. Assess Eval High Educ 2009;34:79–89. doi:10.1080/02602930801895786

13 Tekian A, Watling CJ, Roberts TE, et al. Qualitative and quantitative feedback in the context of competency-based education. Med Teach 2017;39:1245–9. doi:10.1080/0142159X.2017.1372564

14 Harrison CJ, Könings KD, Schuwirth L, et al. Barriers to the uptake and use of feedback in the context of summative assessment. Adv Heal Sci Educ 2015;20:229–45. doi:10.1007/s10459-014-9524-6

15 Watling CJ, Driessen EW, van der Vleuten CPM, et al. Understanding responses to feedback: the potential and limitations of regulatory focus theory. Med Educ 2012;46:593–603. doi:10.1111/j.1365-2923.2012.04209.x

16 Ramani S, Könings KD, Mann K V., et al. About Politeness, Face, and Feedback: Exploring Resident and Faculty Perceptions of How Institutional Feedback Culture Influences Feedback Practices. Acad Med 2018;93:1348–58. doi:10.1097/ACM.0000000000002193

17 van de Ridder JMM, Peters CMM, Stokking KM, et al. Framing of feedback impacts student’s satisfaction, self-efficacy and performance. Adv Heal Sci Educ 2015;20:803–16. doi:10.1007/s10459-014-9567-8

18 Nussbaum AD, Dweck CS. Defensiveness Versus Remediation: Self-Theories and Modes of Self-Esteem Maintenance. Personal Soc Psychol Bull 2008;34:599–612. doi:10.1177/0146167207312960

19 Tweed RG, Lehman DR. Learning Considered within a Cultural Context: Confucian and Socratic Approaches. Am Psychol 2002;57:89–99.https://search.proquest.com/docview/62303086?accountid=12528

20 Harrison CJ, Könings KD, Dannefer EF, et al. Factors influencing students’ receptivity to formative feedback emerging from different assessment cultures. Perspect Med Educ 2016;5:276–84. doi:10.1007/s40037-016-0297-x

21 Suhoyo Y, van Hell EA, Prihatiningsih TS, et al. Exploring cultural differences in feedback processes and perceived instructiveness during clerkships: Replicating a Dutch study in Indonesia. Med Teach 2014;36:223–9. doi:10.3109/0142159X.2013.853117

22 Hofstede G, McCrae RR. Personality and Culture Revisited: Linking Traits and Dimensions of Culture. Cross-Cultural Res 2004;38:52–88. doi:10.1177/1069397103259443

23 Hofstede G. Cultural differences in teaching and learning. Int J Intercult Relations 1986;10:301–20. doi:10.1016/0147-1767(86)90015-5

24 Hofstede GJ. Culture’s consequences: International differences in work-related values. Beverley Hills, CA:: Sage 1980.

25 Hofstede G, Hofstede GJ, Minkov M. Part II: Dimensions of national cultures. In: Cultures and Organizations: Software of the Mind. Mc-Graw-Hill Education 2010. 199.

26 Suhoyo Y, Van Hell EA, Kerdijk W, et al. Influence of feedback characteristics on perceived learning value of feedback in clerkships: does culture matter? BMC Med Educ 2017;17:69. doi:10.1186/s12909-017-0904-5

27 Ratnam-Lim CTL, Tan KHK. Large-scale implementation of formative assessment practices in an examination-oriented culture. Assess Educ Princ Policy Pract 2015;22:61–78. doi:10.1080/0969594X.2014.1001319

28 Ramani S, Post SE, Könings K, et al. “It’s Just Not the Culture”: A Qualitative Study Exploring Residents’ Perceptions of the Impact of Institutional Culture on Feedback. Teach Learn Med 2017;29:153–61. doi:10.1080/10401334.2016.1244014

29 Oxford English Dictionary. ‘culture, n.’ OED Online. June 2014. 2014.http://www.oed.com/view/Entry/45746? (accessed 6 Aug 2014).

30 Hwang A, Ang S, Francesco AM. The silent Chinese: The influence of face and kiasuism on student feedback-seeking behaviors. J Manag Educ 2002;26:70–98. doi:10.1177/105256290202600106

31 Wang T. Understanding Chinese Culture and Learning. In: Australian Association for Research in Education. 2006. 1–14.http://publications.aare.edu.au/06pap/wan06122.pdf

32 Jin L, Cortazzi M. Changing Practices in Chinese Cultures of Learning. Lang Cult Curric 2006;19:5–20. doi:10.1080/07908310608668751

33 Marambe KN, Vermunt JD, Boshuizen HPA. A cross-cultural comparison of student learning patterns in higher education. High Educ 2011;64:299–316. doi:10.1007/s10734-011-9494-z

34 Crozet C. The core tenets of education in ancient India, inspirations for modern times. Int J Pedagog Learn 2012;7:262–5. doi:10.5172/ijpl.2012.7.3.262

35 Kadi W. Education in Islam—Myths and Truths. Comp Educ Rev 2006;50:311–24. doi:10.1086/504818

36 Idrus F. Initiating Culturally Responsive Teaching for Identity Construction in the Malaysian Classrooms. English Lang Teach 2014;7:53–64. doi:10.5539/elt.v7n4p53

37 Fullerton PD. ‘That’s the way we learn’: Exploring influences of culture on medical student learning in Malaysia and Australia (MHPE Thesis - Monash University). 2014.

38 Wong G, Greenhalgh T, Westhorp G, et al. Realist methods in medical education research: what are they and what can they contribute? Med Educ 2012;46:89–96. doi:10.1111/j.1365-2923.2011.04045.x

39 Pawson R, Greenhalgh T, Harvey G, et al. Realist review - a new method of systematic review designed for complex policy interventions. J Health Serv Res Policy 2005;10:21–34. doi:10.1258/1355819054308530

40 Greenhalgh T, Wong G, Westhorp G, et al. Protocol - realist and meta-narrative evidence synthesis: Evolving Standards (RAMESES). BMC Med Res Methodol 2011;11:115. doi:10.1186/1471-2288-11-115

41 Wong G, Greenhalgh T, Westhorp G, et al. RAMESES publication standards: Realist syntheses. J Adv Nurs 2013;69:1005–22. doi:10.1111/jan.12095

42 Pawson R. Digging for nuggets: How ‘bad’ research can yield ‘good’ evidence. Int J Soc Res Methodol Theory Pract 2006;9:127–42. doi:10.1080/13645570600595314

43 Moher D, Shamseer L, Clarke M, et al. Preferred reporting items for systematic review and meta-analysis protocols (PRISMA-P) 2015 statement. Syst Rev 2015;4:1. doi:10.1186/2046-4053-4-1

44 Lockwood C, Porritt K, Munn Z, et al. Chapter 2: Systematic Reviews of Qualitative Evidence. In: JBI Manual for Evidence Synthesis. JBI 2020. Section 2.6.2. doi:10.46658/JBIMES-20-03

45 Veritas Health Innovation. Covidence systematic review software. 2020.http://www.covidence.org

46 Critical Appraisal Skills Programme. CASP Qualitative Checklist. 2018.http://www.casp-uk.net/casp-tools-checklists (accessed 22 Nov 2020).

47 Cook DA, Reed DA. Appraising the Quality of Medical Education Research Methods: The Medical Education Research Study Quality Instrument and the Newcastle-Ottawa Scale-Education. Acad Med 2015;90:1067–76. doi:10.1097/ACM.0000000000000786

48 QSR International. NVIVO Software. 2020.https://www.qsrinternational.com/nvivo-qualitative-data-analysis-software/home

