## Supplementary File-Lit Review Search Strategy for "Culture and understanding the role of feedback for health professions students: Realist synthesis protocol"

**Date of Last Search:** 13 Feb 2020
**Review Name in Covidence:**
Cult & Feedback_2020_Ovid_CINAHL_ERIC

---

**Database: Ovid MEDLINE(R)** <1946 to February 10, 2020>

Search Strategy:

--------------------------------------------------------------------------------

1 Students, Dental/ or Students, Nursing/ or Students, Medical/ or Students, Public Health/ or Students, Health Occupations/ or Students, Pharmacy/ (67487)

2 Clinical Clerkship/ (4968)

3 Education, Medical/ or Education, Nursing/ or Education, Dental/ or Education, Public Health Professional/ or Education, Pharmacy/ (108245)

4 Clinical Competence/ (91249)

5 Faculty, Dental/ or Faculty, Pharmacy/ or Faculty/ or Faculty, Nursing/ or Faculty, Medical/ (34698)

6 1 or 2 or 3 or 4 or 5 (257689)

7 Formative Feedback/ (790)

8 debrief.mp. (272)

9 7 or 8 (1055)

10 Culture/ (32539)

11 Cross-Cultural Comparison/ (25321)

12 Cultural Diversity/ (11270)

13 10 or 11 or 12 (66579)

14 6 and 9 and 13 (2)

15 6 and 13 (3424)

16 Graduate Students/ or International Students/ or Nursing Students/ or Medical Students/ (56080)

17 Learning/ or Student Attitudes/ (63965)

18 Clinical Psychology Internship/ or Medical Internship/ or Internship Programs/ or Clinical Methods Training/ or Professional Supervision/ (0)

19 Teaching Methods/ or Teaching/ (48743)

20 Teachers.mp. [mp=title, abstract, original title, name of substance word, subject heading word, floating sub-heading word, keyword heading word, organism supplementary concept word, protocol supplementary concept word, rare disease supplementary concept word, unique identifier, synonyms] (26332)

21 16 or 17 or 18 or 19 or 20 (173229)

22 FEEDBACK/ (29548)

23 "knowledge of results"/ (972)

24 scaffolding/ (0)

25 experiential learning/ or on the job training/ (28060)

26 22 or 23 or 24 or 25 (58111)

27 "culture (anthropological)"/ or ethnology/ or "racial and ethnic groups"/ or sociocultural factors/ (1582)

28 Cross Cultural Differences/ (0)

29 cultural sensitivity/ or ethnic identity/ or ethnic values/ (0)

30 27 or 28 or 29 (1582)

31 21 and 26 and 30 (0)

32 21 and 26 (7544)

33 6 and 9 (533)

34 32 or 33 (8042)

35 6 and 9 (533)

36 exp Education, Professional/ (293818)

37 exp Students, Health Occupations/ (67629)

38 clerkship*.mp. [mp=title, abstract, original title, name of substance word, subject heading word, floating sub-heading word, keyword heading word, organism supplementary concept word, protocol supplementary concept word, rare disease supplementary concept word, unique identifier, synonyms] (6446)

39 student*.mp. [mp=title, abstract, original title, name of substance word, subject heading word, floating sub-heading word, keyword heading word, organism supplementary concept word, protocol supplementary concept word, rare disease supplementary concept word, unique identifier, synonyms] (262260)

40 trainee*.mp. [mp=title, abstract, original title, name of substance word, subject heading word, floating sub-heading word, keyword heading word, organism supplementary concept word, protocol supplementary concept word, rare disease supplementary concept word, unique identifier, synonyms] (20261)

41 (intern or interns*).mp. [mp=title, abstract, original title, name of substance word, subject heading word, floating sub-heading word, keyword heading word, organism supplementary concept word, protocol supplementary concept word, rare disease supplementary concept word, unique identifier, synonyms] (52870)

42 supervisor*.mp. [mp=title, abstract, original title, name of substance word, subject heading word, floating sub-heading word, keyword heading word, organism supplementary concept word, protocol supplementary concept word, rare disease supplementary concept word, unique identifier, synonyms] (18224)

43 supervisor*.mp. [mp=title, abstract, original title, name of substance word, subject heading word, floating sub-heading word, keyword heading word, organism supplementary concept word, protocol supplementary concept word, rare disease supplementary concept word, unique identifier, synonyms] (18224)

44 36 or 37 or 38 or 39 or 40 or 41 or 42 or 43 (509150)

45 cross-cultural comparison/ (25321)

46 cultural characteristics/ or cultural diversity/ (26880)

47 culture/ (32539)

48 Ethnic Groups/ (60439)

49 transcultur*.mp. [mp=title, abstract, original title, name of substance word, subject heading word, floating sub-heading word, keyword heading word, organism supplementary concept word, protocol supplementary concept word, rare disease supplementary concept word, unique identifier, synonyms] (5056)

50 (cross-cultur* or crosscultur*).mp. [mp=title, abstract, original title, name of substance word, subject heading word, floating sub-heading word, keyword heading word, organism supplementary concept word, protocol supplementary concept word, rare disease supplementary concept word, unique identifier, synonyms] (30637)

51 ethnic*.mp. [mp=title, abstract, original title, name of substance word, subject heading word, floating sub-heading word, keyword heading word, organism supplementary concept word, protocol supplementary concept word, rare disease supplementary concept word, unique identifier, synonyms] (155972)

52 cultural.mp. [mp=title, abstract, original title, name of substance word, subject heading word, floating sub-heading word, keyword heading word, organism supplementary concept word, protocol supplementary concept word, rare disease supplementary concept word, unique identifier, synonyms] (125146)

53 cultures.mp. [mp=title, abstract, original title, name of substance word, subject heading word, floating sub-heading word, keyword heading word, organism supplementary concept word, protocol supplementary concept word, rare disease supplementary concept word, unique identifier, synonyms] (280840)

54 45 or 46 or 48 or 49 or 50 or 51 or 52 or 53 (540281)

55 FORMATIVE FEEDBACK/ (790)

56 (de-brief* or debrief*).mp. [mp=title, abstract, original title, name of substance word, subject heading word, floating sub-heading word, keyword heading word, organism supplementary concept word, protocol supplementary concept word, rare disease supplementary concept word, unique identifier, synonyms] (3257)

57 (feedback or feed* back or fed back).mp. [mp=title, abstract, original title, name of substance word, subject heading word, floating sub-heading word, keyword heading word, organism supplementary concept word, protocol supplementary concept word, rare disease supplementary concept word, unique identifier, synonyms] (128396)

58 (feedforward or feed* forward).mp. [mp=title, abstract, original title, name of substance word, subject heading word, floating sub-heading word, keyword heading word, organism supplementary concept word, protocol supplementary concept word, rare disease supplementary concept word, unique identifier, synonyms] (7583)

59 55 or 56 or 57 or 58 (136088)

60 44 and 54 and 59 (390)

61 "Influence of feedback characteristics on perceived".m_titl. (1)

62 60 and 61 (1)

63 45 or 46 or 47 or 48 or 49 or 50 or 51 or 52 or 53 (556994)

64 44 and 59 and 63 (424)

***************************

**Ovid PsycINFO**

Database: PsycINFO <1806 to February Week 1 2020>

Search Strategy:

--------------------------------------------------------------------------------

1 Graduate Students/ or International Students/ or Nursing Students/ or Medical Students/ (27499)

2 Learning/ or Student Attitudes/ (103225)

3 Clinical Psychology Internship/ or Medical Internship/ or Internship Programs/ or Clinical Methods Training/ or Professional Supervision/ (14277)

4 Teaching Methods/ or Teaching/ (81113)

5 Teachers.mp. [mp=title, abstract, heading word, table of contents, key concepts, original title, tests & measures, mesh] (162804)

6 1 or 2 or 3 or 4 or 5 (325051)

7 FEEDBACK/ (17223)

8 "knowledge of results"/ (798)

9 scaffolding/ (1273)

10 experiential learning/ or on the job training/ (2149)

11 7 or 8 or 9 or 10 (21307)

12 "culture (anthropological)"/ or ethnology/ or "racial and ethnic groups"/ or sociocultural factors/ (74833)

13 Cross Cultural Differences/ (51220)

14 cultural sensitivity/ or ethnic identity/ or ethnic values/ (23447)

15 12 or 13 or 14 (137142)

16 6 and 11 and 15 (65)

***************************

**CINAHL**

S4 S1 AND S2 AND S3 (79)

S3 (MH "Cultural Diversity") OR (MH "Cultural Competence") OR (MH "Ethnological Research") OR (MH "Cultural Values") OR "( culture or cultural ) OR cross cultural OR cultural diversity in education OR ethnic identity OR cultural understanding" (32,860)

S2 (MH "Experiential Learning") OR "feedback in the workplace OR feedback in education OR feed forward OR debriefing for meaningful learning OR experiential learning OR on the job learning (5,858)

S1 ( (MH "Students, Nursing, Graduate") OR (MH "Students, Graduate") OR (MH "Students, Physical Therapy") OR "trainee OR graduate students OR ( learners or students ) OR internship" ) OR ( supervisor OR preceptor OR ( teachers or educators ) OR ( professors or faculty or teachers or instructors ) OR ( lecturer or academic or tutor or faculty or convenor or marker ) ) (362,880)

**************************

**ERIC**

(((MAINSUBJECT.EXACT("Medical Students") OR MAINSUBJECT.EXACT("Premedical Students") OR MAINSUBJECT.EXACT("Graduate Students") OR MAINSUBJECT.EXACT("Foreign Medical Graduates") OR MAINSUBJECT.EXACT("Medical Education") OR MAINSUBJECT.EXACT("Graduates") OR MAINSUBJECT.EXACT("Professional Education") OR MAINSUBJECT.EXACT("Medical Schools")) OR (MAINSUBJECT.EXACT("Practicum Supervision") OR MAINSUBJECT.EXACT("Student Experience") OR MAINSUBJECT.EXACT("Graduate Medical Education") OR MAINSUBJECT.EXACT("Clinical Experience") OR MAINSUBJECT.EXACT("On the Job Training") OR MAINSUBJECT.EXACT("Professional Education")) OR (MAINSUBJECT.EXACT("Clinical Teaching (Health Professions)") OR MAINSUBJECT.EXACT("Higher Education") OR MAINSUBJECT.EXACT("Practicum Supervision") OR MAINSUBJECT.EXACT("Graduate Medical Education") OR MAINSUBJECT.EXACT("Teachers") OR MAINSUBJECT.EXACT("Medical Education") OR MAINSUBJECT.EXACT("Dental Schools") OR MAINSUBJECT.EXACT("Clinical Experience") OR MAINSUBJECT.EXACT("Medical Schools"))) AND PEER(yes)) AND ((MAINSUBJECT.EXACT("Scaffolding (Teaching Technique)") OR MAINSUBJECT.EXACT("Workplace Learning")) OR (MAINSUBJECT.EXACT("Error Correction") OR MAINSUBJECT.EXACT("Questioning Techniques") OR MAINSUBJECT.EXACT("Feedback (Response)") OR MAINSUBJECT.EXACT("Workplace Learning") OR MAINSUBJECT.EXACT("Teacher Response") OR MAINSUBJECT.EXACT("Learning Processes"))) AND ((MAINSUBJECT.EXACT("Cultural Background") OR MAINSUBJECT.EXACT("Cultural Influences") OR MAINSUBJECT.EXACT("Intercultural Communication") OR MAINSUBJECT.EXACT("Cross Cultural Studies") OR MAINSUBJECT.EXACT("Cultural Awareness") OR MAINSUBJECT.EXACT("Asian Culture") OR MAINSUBJECT.EXACT("Cultural Context")) OR

(MAINSUBJECT.EXACT("Cultural Pluralism") OR MAINSUBJECT.EXACT("Ethnology") OR MAINSUBJECT.EXACT("Student Diversity") OR MAINSUBJECT.EXACT("Ethnicity") OR MAINSUBJECT.EXACT("Ethnography")))

Databases: ERIC (212)
